# Preconception Health Research Priorities for Adolescents and Young Adults in Australia

**DOI:** 10.64898/2026.07.18.26358378

**Authors:** Zahra Ali Padhani, Jodie C Avery, Gizachew A Tessema, Keeth L. B. Mayakaduwage, Jacqueline A. Boyle, Danielle Mazza, Sara Ataie, Salima Meherali, Zohra S Lassi

## Abstract

**Background:** Guidelines on pre-pregnancy counselling are primarily clinical, and although recommendations and policy documents on preconception care exist in Australia, they place little or no emphasis on preconception health of adolescents and young adults.

**Objective:** To identify and prioritise unanswered questions and evidence uncertainties concerning preconception health needs of adolescents and young adults residing in Australia.

**Design:** Research priority exercise

**Setting and Participants:** Participants included young interest-holders (18-24 years) and professional interest-holders from the academics, healthcare, policy, community and government sectors residing in Australia.

**Methods:** We followed the James Lind Alliance (JLA) methodology to identify research priorities for preconception health of adolescents and young adults. The process was led by a multidisciplinary steering committee comprising young interest-holders and professional interest-holders (including academics and clinicians). A rapid literature review was conducted from which 80 research questions were developed across ten domains, which were refined through consultation and prioritised via two rounds of online surveys on Qualtrics using a 9-point Likert scale.

**Results:** The participants included 14 young interest-holders in each survey round, with 22 professional interest-holders in the first round and 33 in the second. Participants from across Australia participated in the survey, but most were from South Australia. In the first survey round, 28 questions across seven domains were prioritised by both professional and young interest-holders. This was followed by a reprioritisation exercise, resulting in the final top 10 research questions spanning five domains. The highest-priority research questions identified by the interest-holders concentrated in the domains of violence and mental health; early intervention and prevention; smoking, tobacco, alcohol, and substance use; access to preconception care and the healthcare system; and priority populations.

**Conclusion:** The study identified the top 10 priority research questions informed by professional and young interest-holders. It promotes new research and collaboration while offering guidance on future research investments and on designing preconception interventions for adolescents and young adults in Australia. Turning these priorities into research could improve the health outcomes for adolescents and their future generations.

## INTRODUCTION

Despite recent advances, increasing prenatal care services and research, many individuals are not healthy at the time of conception.^1^ The health prior to conception is associated with maternal, perinatal, and neonatal health outcomes.^1^ Preconception, defined as the period before pregnancy or during active pregnancy planning, offers a window of improvement when individuals of reproductive age have an opportunity to improve their health and health-related behaviours.^2^ Behavioural modifications before pregnancy, such as weight management and lifestyle changes (e.g., increasing physical activity, avoiding alcohol, drug, and tobacco use), can be positive steps to improve the subsequent health of the pregnant person and baby.^3^ Improved preconception health reduces the risk of anaemia, preterm birth, stillbirth, infertility, congenital anomaly, and maternal and child mortality.^4^

According to a recent study, approximately one in four women experiences an unintended pregnancy, with around one-third ending in abortion in Australia.^5^ These pregnancies are often associated with social and structural disadvantage, including younger age, exposure to abuse, poverty, unemployment, lower educational attainment, limited social support, and health system barriers.^5,6^ Preconception knowledge plays a critical role in shaping health behaviours and attitudes towards pregnancy planning.^7^ Increasingly, pregnancy planning is recognised not as a discrete pre-pregnancy event, but as a continuum extending across the life course.^8^ However, recent evidence substantial gaps in knowledge of preconception health and fertility among women, including adolescents.^9-11^ The World Health Organization (WHO) has identified adolescents (10-19 years) as a vulnerable population that has received insufficient targeted attention.^12^ They are more prone to be exposed to risk factors, while also facing barriers to accessing care, engaging in preventive health behaviours, and leaving unsafe environments.^13,14^ Risk-related behaviours during adolescence, including violence, poor diet, substance use, smoking, and unprotected sexual activity, can have significant consequences for both immediate health and future reproductive outcomes.^13,15,16^

Several preconception health frameworks, recommendations and guidelines exist in Australia. However, their implementation remains limited. Currently, there are also no guidelines or targeted strategies for promoting and delivering preconception health interventions to improve access to care, except for the pre-pregnancy counselling guideline for practitioners.^17^ Therefore, access to preconception care remains a major barrier. Other barriers include lack of awareness, affordability and confidentiality issues, and financial constraints.^18,19^ According to the existing evidence, Australian women are more willing and eager to learn about preconception care to adopt healthy behaviours.^19^ A recent qualitative study reported that starting a family was a far-distant notion among adolescents, but they expressed interest in more information on preconception health and fertility.^9^

Evidence suggests that promotion of preconception care can improve health behaviours, including diet, physical activity, and smoking cessation, while also enhancing self-efficacy and knowledge and reducing risk factors, among both men and women, particularly those with chronic medical conditions.^20^ However, given the broad scope of preconception health and limited evidence specific to adolescents and young adults, important research gaps remain. There is a need to identify research uncertainties in this area to support a preventive approach that promotes healthier pregnancies, improves the identification and management of risk factors, and informs the development of interventions that can be delivered across diverse settings, including schools, health services, and community contexts, ultimately improving outcomes for families.

## OBJECTIVE

This study aimed to identify and prioritise unanswered questions and evidence uncertainties concerning the preconception health needs of adolescents and young adults residing in Australia.

## METHODS

A research priority-setting exercise was conducted following the James Lind Alliance (JLA) methodology.^21^ JLA provides a guide for establishing and managing the Priority Setting Partnership (PSP), bringing stakeholders/topic experts and consumers together to identify and prioritise unanswered questions or evidence uncertainties. The JLA-PSP process helps shortlist the top 10 priorities, highlighting important areas of research. These are broad, important areas prioritised by stakeholders and consumers for further research.

In this study, we referred to stakeholders and consumers as interest-holders. Healthcare professionals, academics, clinicians, and policymakers were referred to as professional interest-holders, whereas adolescents and young adults were referred to as young interest-holders. The term interest-holders was recommended by the Multi□Stakeholder Engagement Consortium (MuSE) to replace the term ‘stakeholders.’^22^ It is defined as “groups with legitimate interests in the health issue under consideration.”^22^ The term ‘stakeholders’ was replaced to avoid colonial connotations linked to “staking a claim” on occupied or stolen property from Indigenous People.

Following the JLA methodology, we undertook the following steps to conduct the research priority exercise:

### Step 1: Formation of the Steering Committee

We established a steering committee comprising academics and clinicians with expertise in preconception health and adolescent health in Australia. The committee also included young interest-holders, i.e., individuals aged 18-24 years who have lived in Australia for more than six months. The Committee was responsible for exploring the implications and scope of the PSP exercise, which involved reviewing the broader preconception health domains and the specific research questions that guided the evidence-checking stage.

### Step 2: Conducting a Literature Search and Development of Questions

We conducted a literature search on PubMed and Google Scholar to identify existing research gaps and priorities around the preconception health of adolescents and young adults (10 to 24 years) residing in Australia for more than six months. We reviewed literature between 2010 and 2024, limited to publications in the English language only. We also reviewed studies that targeted women of reproductive age and provided segregated data on adolescents and young adults.

A comprehensive list of potential research priority questions was developed based on the literature search and subsequently categorised into domains to facilitate review and prioritisation. The draft questionnaire was presented and shared with the steering committee on March 17^th^, 2025, to ensure collective agreement on the proposed domains and questions. Through brainstorming sessions, the committee further refined the domains and questions, contributing insights drawn from their expertise and lived experiences. Through structured discussions, we finalised 80 research questions under 10 domains (**Box 1**), thus capturing the nature and focus of each question.

**Box 1: Preconception Health Domains for Adolescents and Young Adults in Australia**

**Domain 1:** Early Intervention and Prevention

**Domain 2:** Access to Preconception Care and the Healthcare System

**Domain 3:** Sexual Reproductive Health (SRH) and Family Planning Services

**Domain 4:** Violence and Mental Health

**Domain 5:** Nutrition, Weight Management, and Physical Activity

**Domain 6:** Smoking, Tobacco, Alcohol, and Substance Use

**Domain 7:** Communicable and Non-communicable Diseases (Including Infections)

**Domain 8:** Exposure to Environment and Chemicals

**Domain 9:** Genetics

**Domain 10:** Priority Populations (including First Nations, culturally and racially marginalised (CARM), and Lesbian, Gay, Bisexual, Transgender, Queer/Questioning, Intersex, and Asexual/Aromantic plus (LGBTQIA+) populations, and people with disability)

### Step 3: Research Prioritisation-Survey 1/Phase I

The initial research priority exercise was conducted through an online survey conducted between 1 June 2025 and 20^th^ September 2025. The survey aimed to gather responses from a representative and diverse group of professional and young interest-holders in Australia. The survey consisted of two parts. The first part included a participant information sheet, consent form, eligibility screening questions, sociodemographic data, and a question on the appropriate timing for initiating preconception care among adolescents and young adults. The second component of the survey focused on a research-priority exercise comprising 80 questions across 10 domains (See **STable 1**). Participants were asked to rate each question on a 1–9 Likert scale, with high (1–3), medium (4–6), or low (7–9) priority. They were also given the option to skip any question.

Separate surveys were developed for professional interest-holders and young interest-holders using the Qualtrics XM software.^23^ The survey of young interest-holders contained questions written in plain language to ensure clarity and ease of understanding. The survey questionnaire was co-designed with the Steering Committee and piloted with a small number of participants. The responses from the pilot test were excluded from the final analysis. The survey was anonymised before distribution. The Steering Committee did not participate in conducting the pilot survey, and the study participants were not members of the Steering Committee.

#### Participants and recruitment

The participants involved in the online ranking of the questions included young interest-holders, i.e., adolescents and young adults (18 to 24 years) residing in Australia for more than six months and professional interest-holders, including healthcare professionals, academics, government officials, and policymakers, who had been working on preconception health or adolescent health in Australia for more than six months.

Professional and young interest-holders were recruited through multiple channels. Study flyers and participant information sheets were disseminated via social media platforms to engage digitally connected professionals and young interest-holders. In addition, materials were shared through WhatsApp groups of Muslim and Christian communities and health organisations, such as Healthy Development Adelaide, Preconception Health Network, SHINE SA, Youth Affairs Council Victoria, Survivors of Torture and Trauma Rehabilitation Services (STTARS), the Consumer and Community Involvement Program (CCI), to facilitate access through trusted local networks. We also engaged the students and advisory group of adolescents and young people at Adelaide University. This multi-channel approach aimed to build trust and ensure the inclusion of both professional and young interest-holders.

To recruit professional interest-holders, we conducted online searches on Google and Google Scholar to identify relevant organisations and individuals, reviewed published literature to identify subject-matter experts, and drew on contacts established through public health conferences. Direct email invitations were sent to identified individuals and organisations. Members of the steering committee further supported recruitment by sharing information within their professional networks. Some participants (professional interest-holders) also connected us with additional experts in the field, helping us maximise our reach. Collectively, these strategies enabled engagement with professional interest-holders across sectors involved in the care and health of adolescents and young adults.

### Step 4: Analysis-Refining Questions After Phase I of Research Prioritisation

After gathering responses from the first survey, all responses were exported from Qualtrics to an Excel sheet, where median scores were calculated for each rated question. This was done separately for responses received from professional interest-holders and young interest-holders. We also calculated the median score for each question by combining responses from both professional and young interest-holders. These scores highlighted uncertainties that were seen as priorities by different groups and where their views aligned or differed. After calculating median scores, all high-prioritised questions (median scores of 1 to ≤ 3) were presented to the Steering Committee on 1^st^ and 16^th^ October 2025 (based on the availability of the steering committee members). The consultation process with the Steering Committee helped us finalise the top 28 questions, which reflected the research priorities of both professional and young interest-holders.

### Step 5: Research Prioritisation – Survey 2/Phase II

Phase II was the final stage of the priority-setting process to prioritise the top 10 research uncertainties identified through consensus on preconception health of adolescents and young adults in Australia.

Following Step 4, the top 28 research priorities were sent to the participants for re-prioritisation. The process of recruitment of participants was similar to Phase I. It was not compulsory for participants to complete the Phase I survey to participate in Phase II. The second survey was conducted between November 10^th^, 2025, and January 30^th^, 2026. This survey also consisted of two parts. The first part included a participant information sheet, consent form, eligibility screening form, and sociodemographic questions. The second component of the survey focused on a research-priority exercise, which consisted of prioritised questions across seven domains.

After gathering responses from participants in the round 2 survey, questions were prioritised as described in Step 4. The top 10 priorities were shared with the steering committee members on February 10^th^, 2026, and their insights and feedback helped prioritise the top 10 unanswered questions about the preconception health of adolescents and young adults residing in Australia.

## RESULTS

In the initial survey, we received 53 responses from professional interest-holders and 44 from young interest-holders. Seven participants from each group were ineligible to participate, and five professional interest-holders and 10 young interest-holders either did not give consent or did not respond to the consent form. Nineteen responses from professional interest-holders and 13 from young interest-holders were incomplete, leaving 22 from professional interest-holders and 14 from young interest-holders in our final sample. In the second round of the survey, we received a total of 63 responses from professional interest-holders and 41 from young interest-holders, of which 17 responses from professional interest-holders and seven from young interest-holders were ineligible. Five young interest-holders either did not give consent or did not respond to the consent form. Sixteen responses from professional interest-holders and 15 from young interest-holders were incomplete, leaving 30 complete responses from professional interest-holders and 14 from young interest-holders in our final sample for the second survey.

In the first round of the survey, most of the professional interest-holders were female professionals from South Australia aged 25-50 years, working in academia with 1-5 years of experience (**Table 1**). Most of the young interest-holders in round 1 were males aged 20-24 years, while in round two we had an equal number of male and female respondents. Of the 22 professional interest-holders, most suggested initiating the intervention during early and late adolescence. On the contrary, most of the young interest-holders reported starting preconception care intervention at young adulthood (20-24 years) or only when planning pregnancy.

**Table 1.**
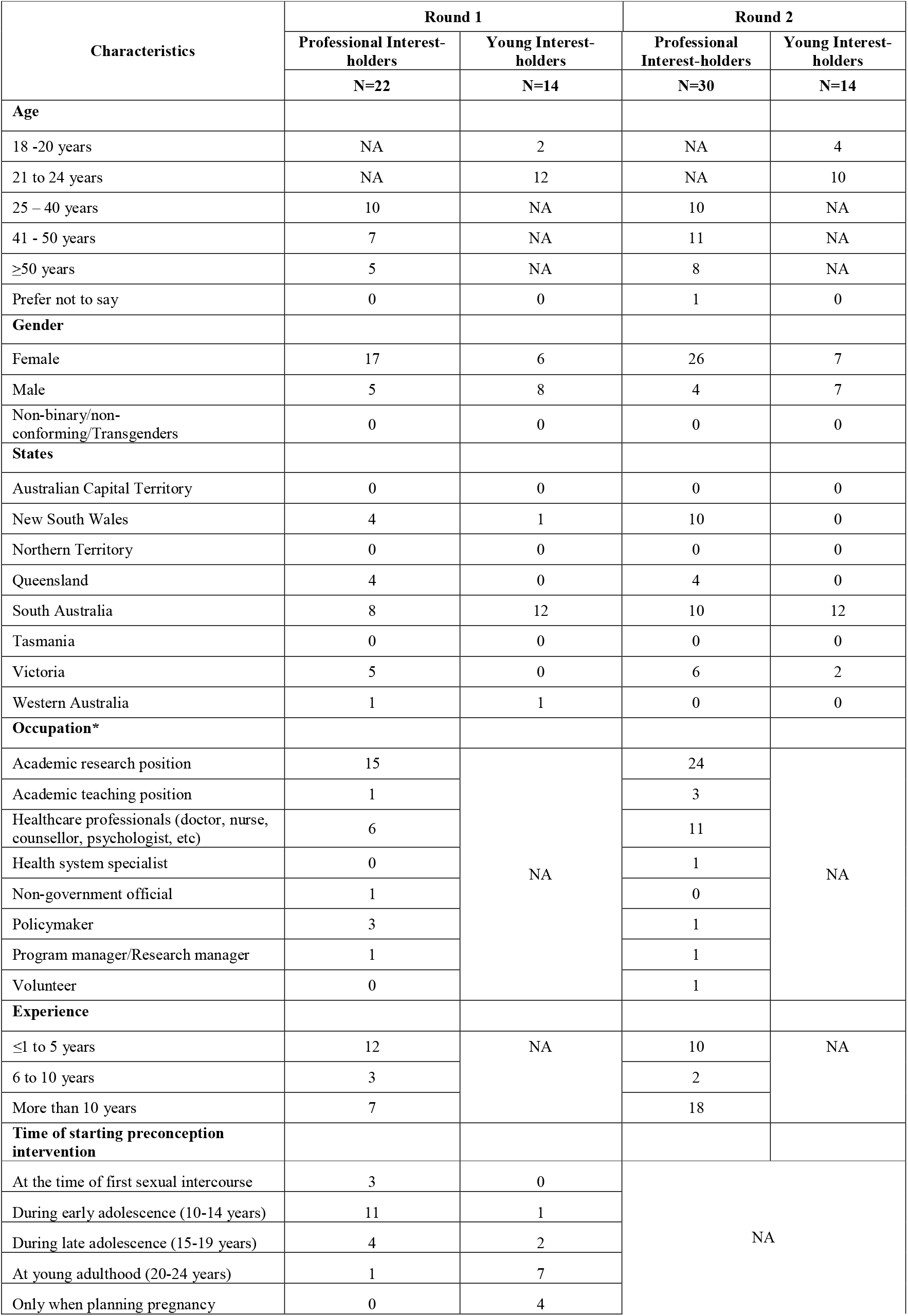
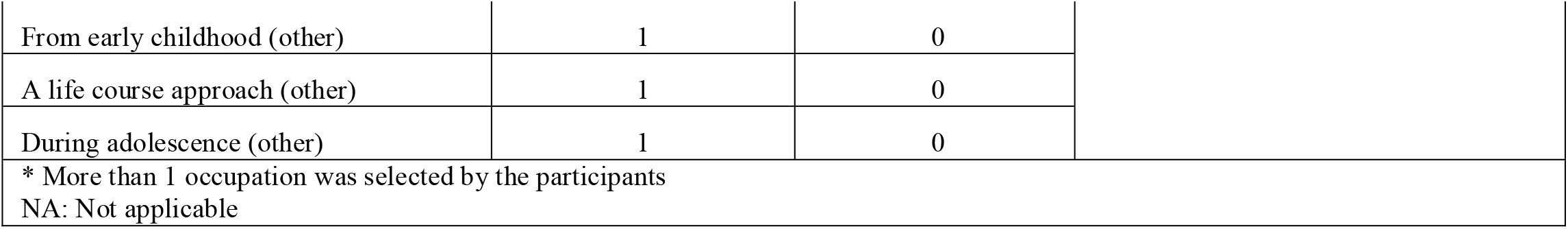
Characteristics of included participants.

| Characteristics | Round 1 |  | Round 2 |  |
| --- | --- | --- | --- | --- |
|  | Professional Interest-holders | Young Interest-holders | Professional Interest-holders | Young Interest-holders |
|  | N=22 | N=14 | N=30 | N=14 |
| **Age** |  |  |  |  |
| 18 -20 years | NA | 2 | NA | 4 |
| 21 to 24 years | NA | 12 | NA | 10 |
| 25 – 40 years | 10 | NA | 10 | NA |
| 41 - 50 years | 7 | NA | 11 | NA |
| ≥50 years | 5 | NA | 8 | NA |
| Prefer not to say | 0 | 0 | 1 | 0 |
| **Gender** |  |  |  |  |
| Female | 17 | 6 | 26 | 7 |
| Male | 5 | 8 | 4 | 7 |
| Non-binary/non-conforming/Transgenders | 0 | 0 | 0 | 0 |
| **States** |  |  |  |  |
| Australian Capital Territory | 0 | 0 | 0 | 0 |
| New South Wales | 4 | 1 | 10 | 0 |
| Northern Territory | 0 | 0 | 0 | 0 |
| Queensland | 4 | 0 | 4 | 0 |
| South Australia | 8 | 12 | 10 | 12 |
| Tasmania | 0 | 0 | 0 | 0 |
| Victoria | 5 | 0 | 6 | 2 |
| Western Australia | 1 | 1 | 0 | 0 |
| **Occupation\*** |  |  |  |  |
| Academic research position | 15 | NA | 24 | NA |
| Academic teaching position | 1 |  | 3 |  |
| Healthcare professionals (doctor, nurse, counsellor, psychologist, etc) | 6 |  | 11 |  |
| Health system specialist | 0 |  | 1 |  |
| Non-government official | 1 |  | 0 |  |
| Policymaker | 3 |  | 1 |  |
| Program manager/Research manager | 1 |  | 1 |  |
| Volunteer | 0 |  | 1 |  |
| **Experience** |  |  |  |  |
| ≤1 to 5 years | 12 | NA | 10 | NA |
| 6 to 10 years | 3 |  | 2 |  |
| More than 10 years | 7 |  | 18 |  |
| **Time of starting preconception intervention** |  |  |  |  |
| At the time of first sexual intercourse | 3 | 0 | NA |  |
| During early adolescence (10-14 years) | 11 | 1 |  |  |
| During late adolescence (15-19 years) | 4 | 2 |  |  |
| At young adulthood (20-24 years) | 1 | 7 |  |  |
| Only when planning pregnancy | 0 | 4 |  |  |

|  |  |  |
| --- | --- | --- |
| From early childhood (other) | 1 | 0 |
| A life course approach (other) | 1 | 0 |
| During adolescence (other) | 1 | 0 |
| * More than 1 occupation was selected by the participants |  |  |
| NA: Not applicable |  |  |

### Survey 1: Initial Prioritisation

Based on the first round of the survey, 24 questions (across five domains) were prioritised. During the steering committee meeting, the members highlighted a concern of excluding priority questions from the other five domains that were not highly prioritised. To address this concern, four more questions that were considered a priority in the combined score (i.e., questions 6.2, 6.4, 7.3 and 7.5) from the unprioritised domains only were included in the list; as a result, 28 questions (across seven domains) were prioritised in the first round of the survey (**Table 2**). Genetics, exposure to the environment and/or chemicals, nutrition, weight management and physical activity domains were not highly prioritised by either professional or young interest-holders.

**Table 2.** Domains and research questions prioritised in the first round of the survey.

| Domains and questions |  |  | Median score |  |  |
| --- | --- | --- | --- | --- | --- |
|  |  |  | Combined | Professional Interest-holders | Young Interest-holders |
| **Domain 1: Early Intervention and Prevention** | 1.1 | What are the preconception risk factors among adolescents and young adults that can help prevent adverse maternal, perinatal, and infant/child health outcomes? | 2.0 | 2.0 | 3.0 |
|  | 1.2 | What are the effective strategies to engage and involve adolescents and young adults in improving their preconception health? | 3.0 | 3.0 | 3.0 |
| **Domain 2: Access to Preconception Care and the Healthcare System** | 2.1 | How can we involve adolescents and young adults in planning, designing, and development of programs to improve preconception health care? | 3.0 | 3.0 | 2.5 |
|  | 2.2 | What strategies can be implemented to enhance the delivery, affordability, and coverage of preconception interventions across various settings, such as urban and rural areas? | 3.0 | 3.0 | 3.0 |
|  | 2.3 | What are the most effective strategies for engaging adolescents and young adults in health education, including school-based programs and online platforms? | 3.0 | 3.0 | 3.0 |
|  | 2.4 | What are the barriers preventing adolescents and young adults from accessing preconception care? | 3.0 | 3.0 | 3.0 |
| **Domain 3: Sexual Reproductive Health (SRH) and Family Planning Services** | 3.1 | What are the long-term health impacts of coercion in reproductive decision-making on perinatal, maternal and child health outcomes? | 3.0 | 3.0 | 3.0 |
|  | 3.2 | To what extent do adolescents and young adults experience coercion in making decisions about pregnancy, such as being pressured to either continue with or terminate a pregnancy? | 2.0 | 3.0 | 2.0 |
|  | 3.3 | What factors contribute to the pressure adolescents and young adults face when deciding whether to keep or end a pregnancy, and how can reproductive health policies and interventions help address these issues? | 3.0 | 3.0 | 3.0 |
|  | 3.4 | What are the existing sexual and reproductive healthcare services/interventions and family planning services available for adolescents and young adults (all genders)? | 3.0 | 3.0 | 3.0 |
|  | 3.5 | What is the knowledge among adolescents and young adults on existing family planning and sexual reproductive health (SRH) services, and the accessibility to it? | 3.0 | 3.0 | 2.5 |
|  | 3.6 | How effective are current school-based SRH programs in improving preconception health knowledge and behaviours? | 2.5 | 3.0 | 2.0 |
| **Domain 4: Violence and Mental Health** | 4.1 | What are the long-term health impacts of violence (be it domestic, sexual, or dating violence) during preconception on perinatal, maternal and child health outcomes? | 3.0 | 3.0 | 3.0 |
|  | 4.2 | What is the impact of mental health issues at preconception on perinatal, maternal and child health outcomes? | 2.0 | 3.0 | 2.0 |
|  | 4.3 | How can the detection and treatment of mental health disorders at preconception help improve pregnancy and child health outcomes? | 3.0 | 3.0 | 2.5 |
|  | 4.4 | How can mental health screening and support be effectively integrated into routine preconception care for adolescents and young adults? | 2.5 | 2.0 | 3.0 |
|  | 4.5 | What strategies could be developed that are effective in preventing intimate partner violence and coerced sex among adolescents and young adults? | 2.0 | 2.0 | 2.5 |
|  | 4.6 | How do programs designed to prevent and reduce gender-based violence equip adolescents and young adults to address and reduce their risk? | 3.0 | 3.0 | 3.0 |
| **Domain 5: Smoking, Tobacco, Alcohol, and Substance Use** | 5.1 | What are the long-term impacts of preconception vaping on pregnancy and the health of future offspring? | 3.0 | 3.0 | 3.5 |
|  | 5.2 | What are the long-term impacts of recreational drug use during preconception on pregnancy and the health of future offspring? | 3.0 | 3.0 | 3.5 |
| **Domain 6: Communicable and Non-communicable Diseases (Including Infections)** | 6.1 | What are the pre-pregnancy predictors of pre-eclampsia/eclampsia among young mothers? | 3.0 | 3.5 | 3.0 |
|  | 6.2 | What are affordable, effective, and feasible ways to screen adolescents and young adults for long-term health issues, for example, high blood pressure, during the preconception period? | 3.0 | 3.0 | 4.0 |
| **Domain 7: Priority Populations** | 7.1 | What are the specific preconception health needs and challenges faced by young migrants and refugees/culturally and racially marginalised (CARM)? | 2.0 | 2.0 | 3.0 |
|  | 7.2 | What are the specific preconception health needs and challenges faced by young First Nations people? | 3.0 | 2.5 | 3.0 |
|  | 7.3 | What are the specific preconception health needs and challenges faced by adolescents and young adults with disabilities? | 2.0 | 2.0 | 3.0 |
|  | 7.4 | What are the specific preconception health needs and challenges faced by young LGBTQIA+ populations? | 3.0 | 2.5 | 3.0 |
|  | 7.5 | How can preconception health services be better tailored to reach and engage First Nations adolescents and young adults? | 2.0 | 2.0 | 2.5 |
|  | 7.6 | How can preconception health services be better tailored to reach and engage adolescents and young adults with a disability? | 2.0 | 2.0 | 3.0 |
| Scoring:<br>• High priority: 1–3 score<br>• Moderate priority: 4–6 score<br>• Low priority: 7–9 score |  |  |  |  |  |

### Survey 2: Prioritising the top 10 research questions

After the second round of survey, the top 10 questions from five domains were prioritised by both professional and young interest-holders, which are presented in **Table 3**. Most of the research questions were from the violence and mental health domain (four questions), followed by the early intervention and prevention domain (two questions), smoking, tobacco, alcohol, and substance use (two questions), access to preconception care and the healthcare system (one question) and the priority populations domain (one question). Communicable and non-communicable diseases (including infections) and sexual reproductive health (SRH) and family planning service domains were prioritised by professional interest-holders, but not by the young interest-holders.

**Table 3.** Domains and research questions prioritised in the second round of the survey.

| Domains and Research Questions |  |  | Median Scores |  |  |
| --- | --- | --- | --- | --- | --- |
|  |  |  | Combined | Professional Interest-holders | Young Interest-holders |
| **Domain 1: Early Intervention and Prevention** | 1.1 | What are the preconception risk factors among adolescents and young adults that can help prevent adverse maternal, perinatal, and infant/child health outcomes? | 3 | 3 | 3 |
|  | 1.2 | What are the effective strategies to engage and involve adolescents and young adults in improving their preconception health? | 2 | 2 | 3 |
| **Domain 2: Access to Preconception Care and the Healthcare System** | 2.4 | What are the barriers preventing adolescents and young adults from accessing preconception care? | 2 | 2.5 | 2 |
| **Domain 4: Violence and Mental Health** | 4.2 | What is the impact of mental health issues at preconception on perinatal, maternal and child health outcomes? | 3 | 3 | 3 |
|  | 4.3 | How can the detection and treatment of mental health disorders at preconception help improve pregnancy and child health outcomes? | 3 | 3 | 3 |
|  | 4.4 | How can mental health screening and support be effectively integrated into routine preconception care for adolescents and young adults? | 3 | 2 | 3 |
|  | 4.5 | What strategies could be developed that are effective in preventing intimate partner violence and coerced sex among adolescents and young adults? | 2 | 2 | 1 |
| **Domain 5: Smoking, Tobacco, Alcohol, and Substance Use** | 5.1 | What are the long-term impacts of preconception vaping on pregnancy and the health of future offspring? | 3 | 3 | 3 |
|  | 5.2 | What are the long-term impacts of recreational drug use during preconception on pregnancy and the health of future offspring? | 3 | 3 | 3 |
| **Domain 7: Priority Populations** | 7.3 | What are the specific preconception health needs and challenges faced by adolescents and young adults with disabilities? | 2.5 | 2 | 3 |
| Scoring:<br>• High priority: 1–3 score<br>• Moderate priority: 4–6 score<br>• Low priority: 7–9 score |  |  |  |  |  |

Of the top 10 questions, the following two were highly prioritised by both professionals and young interest holders, with median scores ranging from 1 to 2.

- *Domain 2: Access to Preconception Care and the Healthcare System*: What are the barriers preventing adolescents and young adults from accessing preconception care?
- *Domain 4: Violence and Mental Health:* What strategies could be developed that are effective in preventing intimate partner violence (IPV) and coerced sex among adolescents and young adults?

## DISCUSSION

The study identified the top 10 research priorities to improve the preconception health of adolescents and young adults in Australia. The highly ranked priorities centred on early intervention and prevention; access to preconception care and health systems; violence and mental health; smoking, tobacco, alcohol and substance use; and preconception health of priority populations, reflecting a shift from traditional biomedical framing toward broader social and behavioural determinants of health. This study also highlights a difference in opinions between professional and young interest-holders regarding the initiation of preconception care. Notably, professional interest-holders recommended initiating preconception interventions in early adolescence, whereas young interest-holders preferred initiating them in young adulthood (20–24 years), suggesting differences in perceived relevance and readiness gap, as adolescents and young adults do not feel ready until their early 20s, while professionals suggest starting early, in middle school or high school. The lack of consensus can also be due to a lack of awareness. A recent study from Australia reported a lack of awareness among the general population regarding preconception care.^24^ Another study reported that only 53% of general practitioners were aware of the preconception care guidelines in Australia.^25^

This study identified a set of shared research priorities but also reported on divergent perspectives among interest-holder groups. Professional interest-holders ranked communicable and non-communicable diseases (including infections) and the sexual and reproductive health and family planning domains as high priorities, but these domains were not considered priorities by the young interest-holders. This difference in perspective reflects the distinct ways in which professionals and young people perceive, experience, and understand the preconception health needs for adolescent and young adults. Nutrition, weight management and physical activity were not considered as a high priority in the first round of the survey by both professional and young interest-holders. Despite their prominence in the preconception health frameworks,^26,27^ these domains were not considered as a high priority by both groups, perhaps highlighting a mismatch with system-level priorities. This misalignment has important implications for the design, acceptability, and uptake of preconception interventions. Genetics, environment and exposure to chemicals were also not prioritised in the first round of the survey; however, they pose a significant threat to maternal and child health. Current evidence also highlights the absence of evidence on environmental or climate exposure on child outcomes, as many studies on pre-pregnancy cohorts have also failed to measure these variables as exposures.^28^

Regarding the priority populations domain, questions on the specific preconception health needs and challenges among the adolescents and young adults with disability emerged in the final top 10 priorities, whereas First Nations, culturally and racially marginalised (CARM), and Lesbian, Gay, Bisexual, Transgender, Queer/Questioning, Intersex, and Asexual/Aromantic plus (LGBTQIA+) populations did not, despite strong prioritisation by professional interest-holders. The variation between professional interest-holders’ scores (consistently high) and young interest-holders’ scores (moderate) is interesting. The difference in how young interest-holders ranked priority populations may reflect their perceived personal relevance rather than a lack of importance. As the demographic information on Indigenous status, ethnicity, or disability was not collected, it is difficult to know whether priorities from these groups were diluted when responses were combined. Importantly, these groups should be viewed through a critical lens that must be integrated across all research activities, rather than as mutually exclusive or topic□specific domains.

Apart from the differences in perspectives of the interest-holder groups, it is important to note the close alignment of violence and mental health, and smoking, tobacco, alcohol, and substance use domains. Several questions within these domains are interconnected, indicating a programmatic or implementation-focused research approach rather than addressing each question in isolation. For example, questions 4.2 to 4.5 from the violence and mental health domain represented a continuum, starting with exposure and impacts to identification, integration into care, and prevention. Similarly, questions 5.1 and 5.2 on the long-term effects of preconception exposure to vaping and recreational drug use from the smoking, tobacco, alcohol, and substance use domain suggest opportunities to be studied together using similar study designs or cohorts.

Lastly, the research priorities centred on violence and mental health; early preconception health intervention and prevention; substance use, access to care, and priority populations. It is not surprising that violence was a high priority area for both interest groups, given the rates of sexual and physical assault experienced by young women.^29^ Exposure to violence or abuse during childhood or before pregnancy has been reported as a strong predictor of family and domestic violence during pregnancy, with poor consequences on maternal and perinatal health outcomes.^30^ According to the current evidence, young Australian women are more at risk of sexual assault compared to older women, resulting in an unwanted pregnancy or termination.^30,31^ A study on pregnant Indigenous women or those with a refugee status in Brisbane, Australia, reported higher rates of IPV compared to non-Indigenous women and those with no refugee status.^32^ Also, those exposed to IPV reported higher odds of pregnancy complications, small for gestational age (SGA) babies, premature birth, and neonatal mortality.^32^ According to existing evidence, COVID-19, social media, financial uncertainty, and political instability have raised levels of distress among young Australians.^33^ This emphasises why both groups prioritised questions related to violence, highlighting the importance of preconception interventions to address violence, especially among adolescents and young adults.

The other identified priorities also align well with the emerging evidence highlighting mental health burden and substance use as key concerns amongst young Australians.^34,35^ The Australian Institute of Health and Welfare reported substance use disorders, mental health, and injuries as major contributors to the high burden of diseases among young Australians aged 15-24 years in 2023.^36^ Similarly, a recent study using the Global Burden of Disease Study 2021 data also reported alcohol and drug use as the leading risk factors of death among adolescents and young adults (10-24 years) in Australia.^35^ Beyond disease burden, these priorities reflect lived experiences shaped by social, economic, and structural factors, including cost-of-living pressures, peer influences, and barriers to accessing care,^34^ underscoring the need for evidence strengthening around preconception health of adolescents and young adults, and emphasising the direction of resources to not only improve their health but the health of families.

The strength of this study is that it captures young interest-holders’ responses, and, to our knowledge, it is the first research priority exercise identifying priority research areas for improving preconception health among adolescents and young adults in Australia. The study has some limitations. First, most responses from the study participants came from South Australia, with very few from other states of Australia. Second, the survey did not collect data on ethnicity, education, First Nations or disability status. Third, several research questions considered as a priority were not considered as a priority by young interest-holders, which may be due to a lack of knowledge and differences in socio-economic and cultural backgrounds. There is also a possibility that some of the groups, such as First Nations, LGBTQIA+, and CARM communities, may be underrepresented.

## CONCLUSION

The study identified the 10 research priorities to improve preconception health of adolescents and young adults in Australia, which centred on violence and mental health; early preconception health intervention and prevention; substance use, access to care, and priority populations. This exercise encourages new research and collaboration, and provides a guide for future research investments, policy development and design of preconception interventions across different settings. Translating these priorities into research has the potential not only to improve the health outcomes of adolescents but also of their future generations.

## Supporting information

Stable 1

## Ethical Approval

Ethical approval was sought from the HREC of the University of Adelaide (now Adelaide University H-2024-176). For this study, participant information sheets and consent forms were provided to study participants at the beginning of the survey. The survey began once the participant agreed to participate. They had the liberty not to answer questions that made them uncomfortable, and to withdraw from the survey at any time.

## Data Availability

All data associated with the study are provided in the manuscript and the supplementary file.

## Acknowledgements

ZSL (#GNT2009730) and GAT (#GNT1195716) are the National Health and Medical Research Council (NHMRC) Investigator Fellows, and JCA is funded via an NHMRC Ideas Grant (#2038988).

## Author contributions

GAT, JCA, ZAP, SM, and ZSL conceptualised the study. DM, KLBM, and JAB were part of the steering committee; ZAP conducted the analysis. All the authors contributed to the interpretation of the results. The first draft of the manuscript was written by ZAP, which was reviewed, edited, and approved by all the authors.

