## Supplementary material for "Preconception Health Research Priorities for Adolescents and Young Adults in Australia": Stable 1

**Stable 1:** Domains and research questions prioritised in the first round of the survey

| **Domains and questions** | | | **Combined** | **Professional Interest-holders only** | **Young Interest-holders** |
| --- | --- | --- | --- | --- | --- |
|  |  |  | **Median** | **Median** | **Median** |
| Domain 1: Early Intervention and Prevention | 1.1 | How do preconception health status and behaviours influence intergenerational health outcomes? | 3.0 | 3.0 | 3.5 |
|  | 1.2 | What is the understanding of preconception health among adolescents and young adults? | 3.0 | 3.0 | 4.0 |
|  | 1.3 | What are the preconception risk factors among adolescents and young adults that can help prevent adverse maternal, perinatal, and infant/child health outcomes? | 2.0 | 2.0 | 3.0 |
|  | 1.4 | When is the best time to offer preconception interventions to support better maternal, perinatal, and infant/child health outcomes? | 3.0 | 3.0 | 3.5 |
|  | 1.5 | What should be included in a core set of preconception health interventions for adolescents and young adults? | 3.0 | 3.0 | 4.5 |
|  | 1.6 | What are the effective strategies to engage and involve adolescents and young adults in improving their preconception health? | 3.0 | 3.0 | 3.0 |
|  | 1.7 | What are the social and economic implications of improving preconception health among adolescents and young adults? | 4.0 | 3.5 | 4.0 |
|  | 1.8 | What indicators should be used to monitor and evaluate preconception health status among adolescents and young adults? | 4.0 | 4.0 | 4.0 |
|  | 1.9 | How data collection methods should be used, and how can they be improved to better track preconception health outcomes among adolescents and young adults? | 4.0 | 4.0 | 5.0 |
| Domain 2: Access to Preconception Care and the Healthcare System | 2.1 | How can adolescent health services be better integrated with maternal care to ensure continuity of care for adolescents who become pregnant and transition into maternal health services? | 3.0 | 3.0 | 3.5 |
|  | 2.2 | How can we involve adolescents and young adults in planning, designing, and development of programs to improve preconception health care? | 3.0 | 3.0 | 2.5 |
|  | 2.3 | What strategies can be implemented to enhance the delivery, affordability, and coverage of preconception interventions across various settings, such as urban and rural areas? | 3.0 | 3.0 | 3.0 |
|  | 2.4 | What are the most effective strategies for engaging adolescents and young adults in health education, including school-based programs and online platforms? | 3.0 | 3.0 | 3.0 |
|  | 2.5 | What are the barriers preventing adolescents and young adults from accessing preconception care? | 3.0 | 3.0 | 3.0 |
|  | 2.6 | What role can digital health play in improving access to preconception care? | 4.0 | 3.5 | 4.5 |
|  | 2.7 | What are the feasibility and costs of implementing digital and other health information technologies to enhance preconception health? What are the key barriers to implementing these technologies? | 4.5 | 4.0 | 5.0 |
|  | 2.8 | What are the harmful impacts of digital platforms on adolescents and young adults, and how can these platforms be used to prevent negative behaviours or health outcomes? | 4.0 | 4.0 | 4.5 |
|  | 2.9 | What role do Participatory Learning and Action (PLA) interventions play in improving access to health services for adolescents and young adults? (Note: PLA is a participatory methodology where community members are actively involved in analysing their own situations, learning together, and taking action to address problems [Ref: Gosling and Edwards 2003]) | 4.0 | 4.0 | 4.0 |
|  | 2.10 | How effective are Participatory Learning and Action (PLA) interventions in improving adolescent and young adult access to healthcare, and what are the key barriers to their implementation? (Note: PLA is a participatory methodology where community members are actively involved in analysing their own situations, learning together, and taking action to address problems [Ref: Gosling and Edwards 2003]) | 4.0 | 4.5 | 3.0 |
| Domain 3: Sexual Reproductive Health and Family Planning Service | 3.1 | What are the long-term health impacts of coercion in reproductive decision-making on perinatal, maternal and child health outcomes? | 3.0 | 3.0 | 3.0 |
|  | 3.2 | To what extent do adolescents and young adults experience coercion in making decisions about pregnancy, such as being pressured to either continue with or terminate a pregnancy? | 2.0 | 3.0 | 2.0 |
|  | 3.3 | What factors contribute to the pressure adolescents and young adults face when deciding whether to keep or end a pregnancy, and how can reproductive health policies and interventions help address these issues? | 3.0 | 3.0 | 3.0 |
|  | 3.4 | What are the practices of female genital mutilation, and how do they impact maternal, perinatal, and child health outcomes | 4.0 | 4.0 | 4.0 |
|  | 3.5 | What are the existing sexual and reproductive healthcare services/interventions and family planning services available for adolescents and young adults (all genders)? | 3.0 | 3.0 | 3.0 |
|  | 3.6 | What is the knowledge among adolescents and young adults on existing family planning and sexual reproductive health (SRH) services and the accessibility to it? | 3.0 | 3.0 | 2.5 |
|  | 3.7 | How do migration status, cultural identity and norms impact the SRH experiences and decision-making behaviours of adolescents and young adults (of all genders)? | 3.0 | 2.5 | 3.5 |
|  | 3.8 | What are the family planning methods utilised by adolescents and young adults (all genders) to prevent unintended pregnancies? | 3.5 | 3.5 | 3.5 |
|  | 3.9 | What are the family planning methods utilised by adolescents and young adults (all genders) to prevent sexually transmitted diseases/infections? | 4.0 | 4.0 | 3.5 |
|  | 3.10 | What are the experiences and preferences of adolescents and young adults regarding contraceptive methods and decision-making? | 4.0 | 3.5 | 5.5 |
|  | 3.11 | What are the perceptions and attitudes of parents/guardians towards family planning and contraception for their children? | 4.0 | 4.0 | 4.0 |
|  | 3.12 | What approaches can be developed and adopted to increase awareness and improve access to emergency contraception?? | 3.5 | 3.5 | 3.5 |
|  | 3.13 | Which community-based programs (for example: youth mentoring programs) can help improve SRH outcomes among females? | 3.5 | 3.0 | 4.5 |
|  | 3.14 | Which community-based programs (for example: youth mentoring programs) can help improve SRH outcomes among males? | 3.0 | 3.0 | 4.5 |
|  | 3.15 | Which community-based programs (for example: youth mentoring programs) can help improve SRH outcomes among the LGBTQIA+ population? | 3.0 | 3.0 | 3.5 |
|  | 3.16 | How effective are current school-based sexual and reproductive health programs in improving preconception health knowledge and behaviours? | 2.5 | 3.0 | 2.0 |
| Domain 4: Violence and Mental Health | 4.1 | What are the long-term health impacts of violence (be it domestic, sexual, or dating violence) during preconception on perinatal, maternal and child health outcomes? | 3.0 | 3.0 | 3.0 |
|  | 4.2 | What is the impact of mental health issues at preconception on perinatal, maternal and child health outcomes? | 2.0 | 3.0 | 2.0 |
|  | 4.3 | How does the use of anti-depressants and anti-anxiety medications during the preconception period impact maternal, perinatal, and child health outcomes? | 3.0 | 4.0 | 3.0 |
|  | 4.4 | How can the detection and treatment of mental health disorders at preconception help improve pregnancy and child health outcomes? | 3.0 | 3.0 | 2.5 |
|  | 4.5 | How can mental health screening and support be effectively integrated into routine preconception care for adolescents and young adults? | 2.5 | 2.0 | 3.0 |
|  | 4.6 | How can community health workers effectively and feasibly deliver a preventive mental health intervention package, with linkages to the primary health care system for treatment? | 3.0 | 3.0 | 4.0 |
|  | 4.7 | What strategies could be developed that are effective in preventing intimate partner violence and coerced sex among adolescents and young adults? | 2.0 | 2.0 | 2.5 |
|  | 4.8 | How do programs designed to prevent and reduce gender-based violence equip adolescents and young adults to address and reduce their risk? | 3.0 | 3.0 | 3.0 |
| Domain 5: Nutrition, Weight Management, and Physical Activity | 5.1 | What is the impact of preconception weight (underweight, overweight or obesity) and lifestyle during adolescence on maternal and perinatal health outcomes? | 4.0 | 4.0 | 4.5 |
|  | 5.2 | What is the impact of preconception weight (underweight, overweight or obesity) and lifestyle during adolescence on the growth and development of their child? | 5.0 | 5.0 | 6.0 |
|  | 5.3 | What is the impact of preconception dietary patterns and physical activity on pregnancy and birth outcomes? | 4.0 | 5.0 | 3.5 |
|  | 5.4 | What is the impact of preconception dietary quality during adolescence on pregnancy and birth outcomes? | 4.0 | 4.0 | 3.5 |
|  | 5.5 | What are the existing nutrition interventions targeted at adolescents and young adults during preconception to improve their maternal, perinatal, and child health outcomes? | 4.0 | 4.0 | 5.0 |
| Domain 6: Smoking, Tobacco, Alcohol, and Substance Use | 6.1 | What are the long-term effects of preconception exposure to tobacco smoke on pregnancy and the health of future offspring? | 4.0 | 5.0 | 3.0 |
|  | 6.2 | What are the long-term impacts of preconception vaping on pregnancy and the health of future offspring? | 3.0 | 3.0 | 3.5 |
|  | 6.3 | What are the long-term impacts of preconception alcohol use on pregnancy and the health of future offspring? | 4.0 | 5.0 | 4.0 |
|  | 6.4 | What are the long-term impacts of recreational drug use during preconception on pregnancy and the health of future offspring? | 3.0 | 3.0 | 3.5 |
|  | 6.5 | What is the impact of preconception interventions to prevent smoking among adolescents and young adults on pregnancy outcomes? | 4.0 | 4.0 | 4.0 |
|  | 6.6 | What is the impact of preconception interventions to prevent vaping among adolescents and young adults on pregnancy outcomes? | 4.0 | 2.0 | 5.0 |
|  | 6.7 | What is the impact of preconception interventions to prevent alcohol use among adolescents and young adults and its impact on pregnancy outcomes? | 4.0 | 4.0 | 3.5 |
|  | 6.8 | What is the impact of preconception interventions to prevent recreational drug use among adolescents and young adults and its impact on pregnancy outcomes? | 4.0 | 3.0 | 4.0 |
| Domain 7: Communicable and Non-communicable Diseases (Including Infections) | 7.1 | What are the pre-pregnancy predictors of gestational diabetes among young mothers? | 4.0 | 4.0 | 3.0 |
|  | 7.2 | What are the pre-pregnancy predictors of gestational hypertension among young mothers? | 4.0 | 4.0 | 4.0 |
|  | 7.3 | What are the pre-pregnancy predictors of pre-eclampsia/eclampsia among young mothers? | 3.0 | 3.5 | 3.0 |
|  | 7.4 | How does the use of medications for conditions like epilepsy or cardiac problems during the preconception period affect maternal, perinatal, and child health outcomes? | 4.0 | 4.0 | 4.0 |
|  | 7.5 | What are affordable, effective, and feasible ways to screen adolescents and young adults for long-term health issues, for example, high blood pressure, during the preconception period? | 3.0 | 3.0 | 4.0 |
|  | 7.6 | What are affordable, effective, and feasible ways to screen adolescents and young adults for infections during the preconception period? | 4.0 | 4.0 | 4.5 |
|  | 7.7 | What strategies are available to improve STI/HIV prevention and management as part of preconception care? | 4.0 | 4.0 | 4.0 |
|  | 7.8 | How does HPV vaccination during adolescence affect maternal, perinatal, and childhood health outcomes?? | 4.5 | 4.0 | 5.5 |
| Domain 8: Exposure to Environment and Chemicals | 8.1 | What is the impact of environmental and chemical exposures during preconception on maternal, perinatal and child health? | 4.0 | 4.5 | 4.0 |
|  | 8.2 | What is the impact of preconception occupational exposures to cleaning products and disinfectants on maternal, perinatal and child health outcomes? | 4.0 | 4.0 | 4.0 |
|  | 8.3 | What public health policies and strategies are in place to regulate and reduce environmental exposures and harmful chemicals? | 4.0 | 5.0 | 4.0 |
|  | 8.4 | What strategies or approaches can help modify individuals' behaviour to reduce environmental exposure to prevent long-term health impacts? | 4.0 | 4.5 | 4.0 |
| Domain 9: Genetics | 9.1 | What are the preconception genetic risk factors among adolescents and young adults that can affect pregnancy and child health outcomes | 5.0 | 5.0 | 4.5 |
|  | 9.2 | What is the role of genetic testing during preconception and its impact on pregnancy and child health outcomes? | 5.0 | 5.0 | 4.5 |
|  | 9.3 | How aware are adolescents and young adults of genetic screening options and genetic counselling, and how accessible are these services to them, especially in relation to preconception health? | 4.0 | 4.0 | 4.5 |
|  | 9.4 | What is the feasibility and cost-effectiveness of implementing community-based genetic screening programs? | 5.0 | 5.0 | 4.5 |
| Domain 10: Priority Populations | 10.1 | What are the specific preconception health needs and challenges faced by young migrants and refugees/culturally and racially marginalised (CARM)? | 2.0 | 2.0 | 3.0 |
|  | 10.2 | What are the specific preconception health needs and challenges faced by young First Nations people? | 3.0 | 2.5 | 3.0 |
|  | 10.3 | What are the specific preconception health needs and challenges faced by adolescents and young adults with disabilities? | 2.0 | 2.0 | 3.0 |
|  | 10.4 | What are the specific preconception health needs and challenges faced by young LGBTQIA+ populations? | 3.0 | 2.5 | 3.0 |
|  | 10.5 | How can preconception health services be better tailored to reach and engage adolescents and young adults from CARM, and migrant and refugee backgrounds? | 2.0 | 2.0 | 4.0 |
|  | 10.6 | How can preconception health services be better tailored to reach and engage First Nations adolescents and young adults? | 2.0 | 2.0 | 2.5 |
|  | 10.7 | How can preconception health services be better tailored to reach and engage adolescents and young adults with a disability? | 2.0 | 2.0 | 3.0 |
|  | 10.8 | How can preconception health services be better tailored to reach and engage young LGBTQIA+ populations? | 2.0 | 2.0 | 3.5 |
| Scoring:   - High priority: 1–3 score - Moderate priority:4–6 score - Low priority: 7–9 score | | | | | |
